# Sewage surveillance for the presence of SARS-CoV-2 genome as a useful wastewater based epidemiology (WBE) tracking tool in India

**DOI:** 10.1101/2020.06.18.20135277

**Authors:** Sudipti Arora, Aditi Nag, Jasmine Sethi, Jayana Rajvanshi, Sonika Saxena, Sandeep K Shrivastava, A. B. Gupta

## Abstract

The infection with SARS-CoV-2 is reported to be accompanied by the shedding of the virus in stool samples of infected patients. Earlier reports have suggested that COVID-19 agents can be present in the fecal and sewage samples and thus it can be a good indication of the pandemic extent in a community. However, no such studies have been reported in the Indian context so far. Since, several factors like local population physiology, the climatic conditions, sewage composition, and processing of samples could possibly affect the detection of the viral genome, it becomes absolutely necessary to check for the presence of the SARS-CoV-2 in the wastewater samples from wastewater treatment plants (WWTPs) serving different localities of Jaipur city, which has been under red zone (pandemic hotspots) since early April 2020. Samples from different local municipal WWTPs and hospital wastewater samples were collected and wastewater based epidemiology (WBE) studies for the presence of SARS-CoV-2 were carried out using the RT-PCR technique to confirm the presence of different COVID-19 target genes namely S gene, E gene, ORF1ab gene, RdRp gene and N gene in the viral load of wastewater samples. In the present study, the untreated wastewater samples from the municipal WWTPs and hospital wastewater samples showed the presence of SARS-CoV-2 viral genome, which was correlated with the increased number of COVID-19 positive patients from the concerned areas, as per reported in the publically available health data. This is the first study that investigated the presence of SARS-CoV-2 viral genome in wastewater, at higher ambient temperature (above 40°C), further validating WBE as a potential tool in predicting and mitigating outbreaks.

**Highlights:**

- The study reports detection of SARS-CoV-2 in sewage in India.
- The presence of SARS-CoV-2 was confirmed by RT-PCR.
- The presence of viral genome was detected at high ambient temperatures of 40-45° C.
- Corroborates trends in the WWTPs showing viral genome with public health data.
- Treated effluent from WWTPs appears safe for reuse with low public health concern.

**Graphical Abstract:** 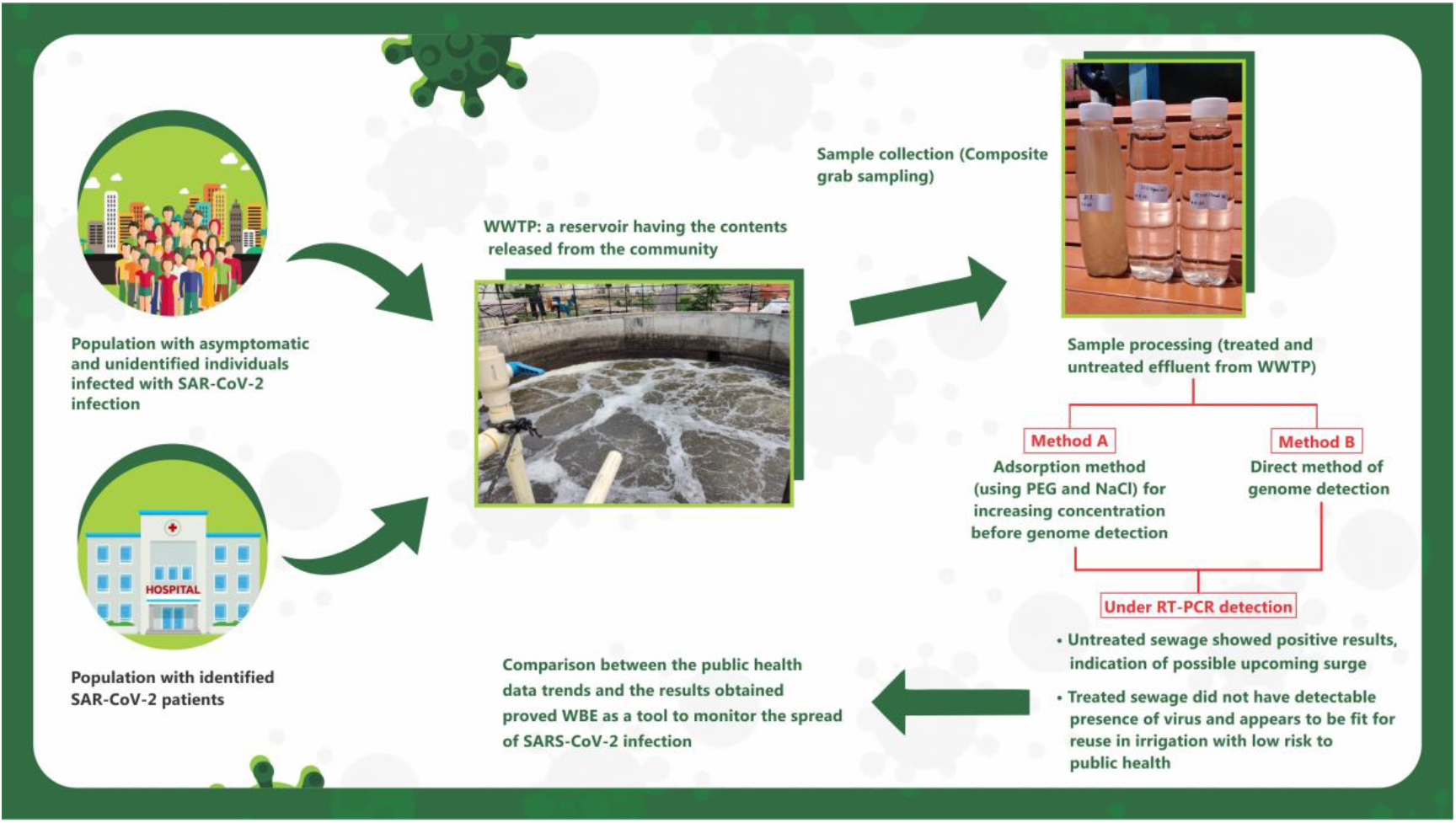

## 1. Introduction

Coronavirus can be considered as a paradigm of the phenomenon involving complex dynamics of animals, humans and environments and in the last 18 years, it has caused three new alarming diseases: severe acute respiratory syndrome (SARS), Middle East respiratory syndrome (MERS), and the current coronavirus disease 2019 (COVID-19) (WHO, 2020a). The Outbreak of zoonotic coronavirus in humans was first reported in Wuhan, China in 2019 as a pneumonia-like ailment caused by a hitherto uncharacterized etiologic agent (Casanova, et al., 2020). The Coronavirus study group of the International Committee of taxonomy of viruses later classified this zoonotic virus as SARS-CoV-2 based on the features it exhibited. (WHO, 2020b). Researchers classified this zoonotic agent into the Coronaviridae family on the basis of its genetic structure and crown-like (or halo) structures present on its envelope glycoprotein and characteristic features of chemistry and replication (Tyrrell DAJ & Myint SH, 1996). The affliction of this single-stranded positive-sense RNA virus shows varying symptoms in individuals depending upon their immune system. According to the data compiled by WHO, some of the commonly reported symptoms include fever, dry cough, tiredness and in some cases may also manifest in aches and pains, nasal congestion, sore throat or diarrhea, conjunctivitis, headache, loss of taste or smell, a rash on the skin, or discoloration of fingers or toes. Serious symptoms reported include difficulty breathing or shortness of breath, chest pain or pressure, loss of speech or movement (WHO 2020c).

The novel strain of coronavirus has evolved after mutations in the previously existing strains. It has been reported that animals played a key role in spreading coronaviruses in humans until recently. But, the mutations, which led to the evolution of SARS-CoV-2, have equipped it with the ability to transmit directly from one human to another (Chan et al., 2020). COVID-19 is reported to transmit even through the droplets from an infected person’s sneeze or even breath (WHO2020b, Chan et al., 2020). Both viable SARS-CoV-2 and viral RNA are shed in bodily excreta, including saliva, sputum, and feces, which are subsequently found in wastewater (Wu et al., 2020a). It is believed that the major transmission route of this virus is inhalation via person-to-person and aerosol/droplet transmission or through fomite, and close contact. At present, COVID-19 is responsible for a rapidly expanding global epidemic with tens of thousands of cases and thousands of deaths (Heymann and Shindo, 2020). The potential fecal-oral transmission was recently highlighted by Yeo et al., (2020). It is therefore possible that the virus may be released with wastewater and contaminates other water bodies (surface, sea, and groundwater), generating aerosols. Sewage from hospitals, especially infectious disease units, may contain the epidemic virus, thus requiring efficient disinfection before discharge into natural waters. Currently, available evidence indicates the need for a better understanding of the role of wastewater as potential sources of epidemiological data and as a factor in public health risk.

Environmental Surveillance is a tool used to monitor the extent and duration of the spread of the virus in specific populations. It gives a measure of contaminants and also provides warning of possible threats emerging in that particular confinement. These monitoring tools have already been successfully implemented for viruses such as poliovirus and the Aichi virus, in the past. The study of wastewater based epidemiology has been proven successful in other cases as well such as Influenza A (H1N1) epidemic 2009 (Heijnen & Medema, 2011), Norovirus, Hepatitis A virus, Hepatitis E virus, Adenovirus, Astrovirus and Rotavirus (Hellmér et al., 2014) in determining the viral concentration in the sewage sample both before and after the onset of the symptoms, which have aided in strategies that resulted in their elimination. This technique is in line with the wastewater based epidemiology (WBE) concept. WBE is a reliable surveillance model for identifying global hotspots of COVID-19 (Núñez-Delgado, A. et al., 2020). Crucially, according to a recent study, SARS-CoV-2 RNA was detected in samples of sewage before any case was reported, suggesting that virus monitoring could be feasible before cases are documented through the health surveillance system (Orive, G., et al., 2020).

Human CoVs, including SARS-CoV and MERS-CoV, are known to cause gastrointestinal symptoms as well as respiratory symptoms. In fact, previous studies demonstrated that these viruses replicate in the gastrointestinal tract (Leung et al., 2003; Zhou et al., 2017). Recent reports revealed that 2–10% of COVID-19 patients had gastrointestinal symptoms, including diarrhea (Chen et al., 2020; Wang et al., 2020a). Although the exact mechanism of COVID-19-induced gastrointestinal symptoms largely remains unknown, a recent study reported that SARS-CoV-2 infects gastrointestinal glandular epithelial cells (Wu et al., 2020b). With diarrhea being reported as a frequent symptom in the patient (Kitajima, et al., 2020; D’Amico, et al., 2020), the shedding of SAR-CoV-2 RNA through feces has been reported in many countries like the Netherlands (Medema et al., 2020), Wuhan, China (Chen et al., 2020; Hindson, 2020). A study conducted on the clinical samples collected from different hospitals in China reveals shedding of viral RNA through feces in a significant number of cases, implying the fecal route of transmission (Wang et al., 2020a). Some clinical studies reported prolonged fecal shedding of SARS-CoV-2 RNA for up to seven weeks after the first symptom onset (Wu et al., 2020a). Another study reported that viral RNA could be detected in the feces of 81.8% cases even with a negative throat swab result (Ling et al., 2020). Recent reports implied that significant proportions (17.9–30.8%) of infected individuals are asymptomatic (Wu, et al., 2020b; Tang et al., 2020). It will be easier to survey regions for viral infections, especially in the asymptomatic cases of COVID-19, comprehensively and in real-time for the reason that the affected individuals start shedding the viral RNA genomes in their feces. This tool will serve as an efficient and sensitive monitoring tool to measure virus levels in the hotspot populations and provide early warning signs before a potential epidemic in the future. The current approach analyzes wastewater samples to determine the presence of infected individuals and estimate the number of cases. As of May 7, 2020, India counted 52,952 cases out of which 35, 902 were active and it showed a tremendous increase by 20.56% and 28.39% in total and active cases, respectively, within a month. As of June 7, 2020, this number was reported to be 257,486 total and 126,431 active cases, respectively (official data from John Hopkins University, 2020). The data from the sewage supports communities with trend analysis that can determine the pandemic hotspots of COVID-19. This further can help researchers to generate early warning and thus help officials to take rapid action regarding the containment or re-emergence of new outbreaks in a population.

In this context, the present study was planned to achieve the evidence for detection of SARS-CoV-2 RNA samples in Municipal WWTPs for untreated and treated wastewater samples, and hospital sewage samples around Jaipur city, and determine the correlations between the positive results for sewage samples from WWTPs with the public health data, as officially reported in the daily newspaper, of positive patients around the area. To date, there have been no reports of detection of SARS-CoV-2 in wastewater in India. The present study provides the first reported evidence of the presence of SARS-CoV-2 RNA in sewage samples of Jaipur, Rajasthan (India) and these findings demonstrate the applicability of WBE or sewage surveillance as an early indicator of persistence of the virus in the community and the risk associated with wastewater handling. The work also draws attention to the current wastewater treatment system being used in WWTPs, and the efficacy of sodium hypochlorite or other chlorine compounds being used by hospital authorities as a potent disinfecting agent, to inactivate or attenuate viruses.

## 2. Experimental Methodology

### 2.1. Wastewater sampling

Wastewater samples were collected from different units of six municipal wastewater treatment plants (WWTPs) installed at different locations of Jaipur city and wastewater samples from two hospitals, which are the major treatment centers for COVID-19 patients. The location of 8 sites (six municipal WWTPS and 2 Hospitals) is shown in Figure 1. The samples were taken between 3^rd^ May 2020 and 14^th^ June 2020. All the samples were collected in sterile bottles and transported to the Environmental Biotechnology laboratory at Dr. B. Lal Institute of Biotechnology, Jaipur for further investigations. Sample collection was carried out by taking appropriate precautions with the use of standard personal protective equipment (PPE). Table 1 highlights the comprehensive list of all the sample locations along with dates, types of samples collected, and current treatment technology.

**Table 1:**
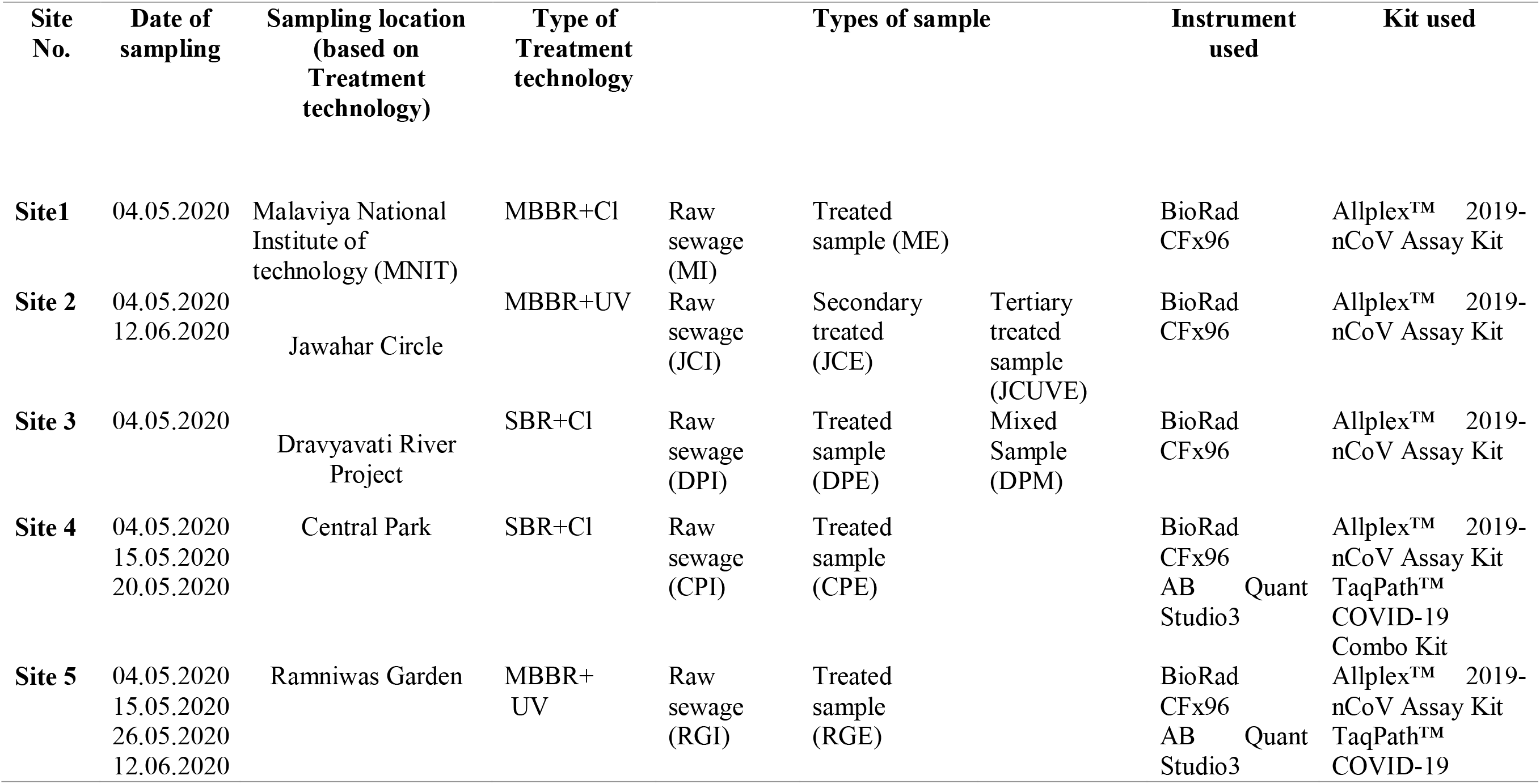
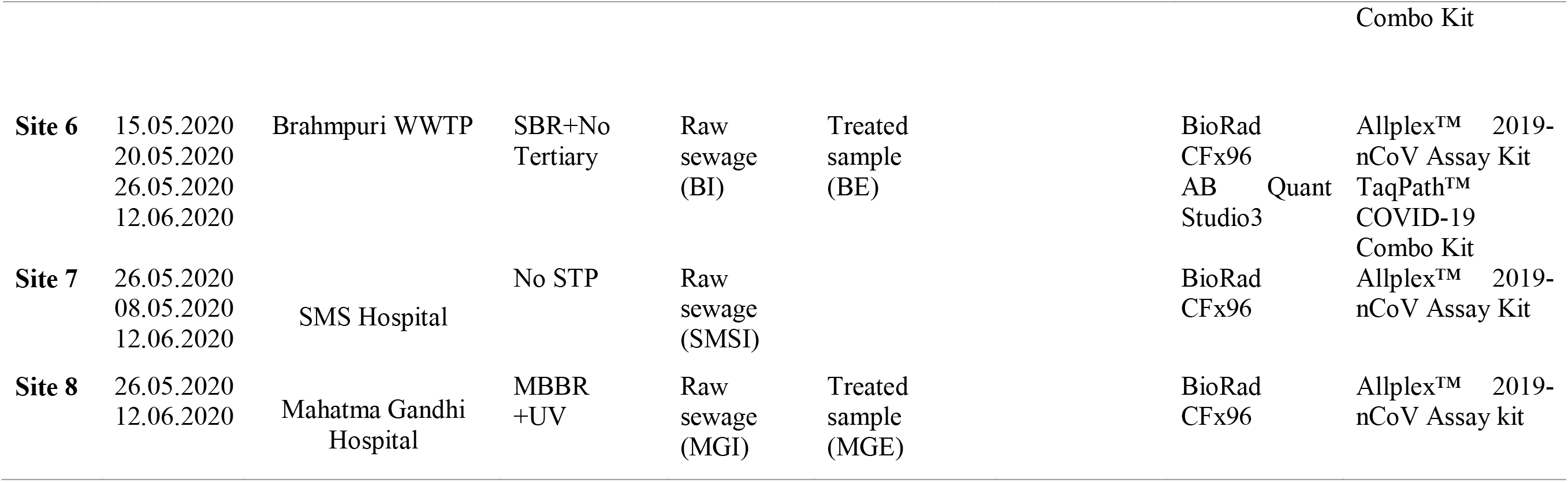
Types of samples and sampling locations.

**Figure 1:**
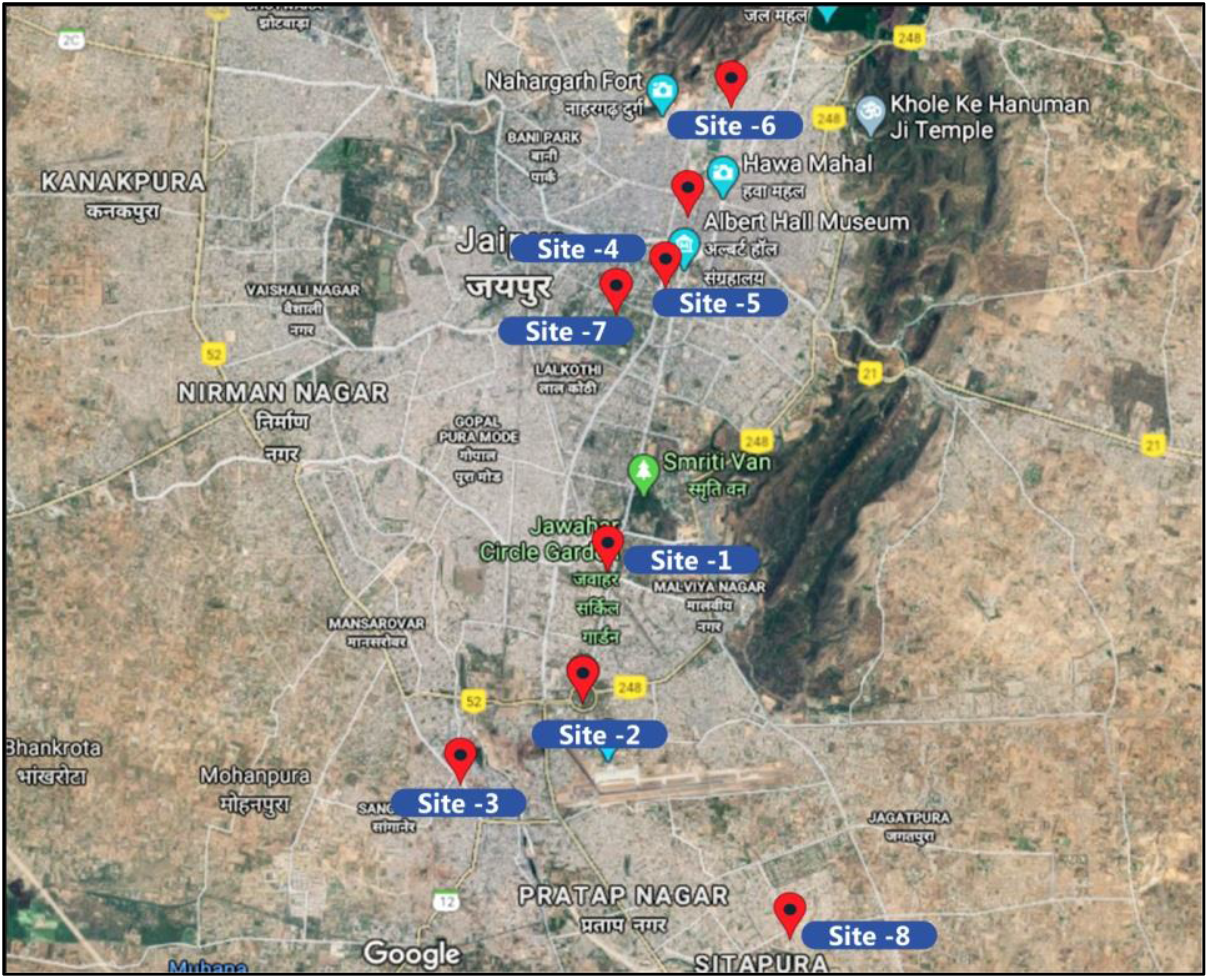
Map showing locations of sample collection from Jaipur (India) *as shown by red pins*. Sites 1-6 are Municipal WWTPs, and Site 7 and 8 are Hospitals (SMS Hospital & Mahatma Gandhi Hospital WWTP)

### 2.2. Sample Pre-Processing

Samples were processed using two different methods based on previously published protocols (La Rosa, et al., 2020, Biobot Analytics). The two methods were tested and standardized for sample pre-processing in the laboratory. In the first method, (Method A), sample processing was carried out in two different steps involving the inactivation of virus particles (which might be present in the wastewater sample), followed by the concentration of the virus by adsorption. Method A began with inactivation of the virus by transferring the samples to 50 ml Tarsons falcon (code 546041) in a biosafety cabinet (BSL2), followed by 70% ethanol spray over the surface of the falcon tubes and UV light exposure for 30 minutes for surface sterilization. After UV exposure, the samples were transferred to the water bath at 60°C and kept incubated for 90 minutes for heat inactivation of the virus. After the inactivation of the coronavirus, the samples were brought to room temperature and filtered through a 0.45 μm membrane using a vacuum filter assembly. The filtrate of each respective sample was transferred to a fresh 50 ml falcon containing 0.9 g sodium chloride (NaCl) and 4 g polyethylene glycol (PEG). The contents were dissolved by gentle manual mixing. The samples containing PEG and NaCl were then centrifuged at 4 °C for 30 minutes at 7000 rpm. The pellet obtained was further re-suspended in 1X Phosphate Buffer Saline (PBS). In the second method, which is a direct method (Method B), for detection of SARS-CoV-2 in the wastewater sample includes UV treatment of the sample for 30 minutes, followed by dispensing 1 ml sample in each 1.7 ml centrifuge tube. A spin on 7,000 rpm for 15 minutes was given to each tube containing the samples. The supernatant was collected in a separate tube and the same process was repeated. The supernatant thus obtained was then processed for RNA extraction.

### 2.3. RNA extraction

RNA extraction and subsequent steps of detection were done at Dr. B. Lal Clinical Laboratory Pvt. Ltd, Jaipur (which is authorized by ICMR to conduct COVID-19 testing in humans). For the extraction of viral RNA, FDA and ICMR approved Allplex™ 2019-nCoV Assay kit (cat# RP10244Y RP10243X) was used. 10 μl of proteinase K and 200 μl of lysis buffer were added to 200 μl of the sample into a 1.5 ml centrifuge tube followed by vortex mixing and incubation at 56°C for 15 minutes in a heating block. 250 μl of ethanol was added to the sample and mixed by pulse vortexing for 15 seconds. The mixture was then transferred to the spin column and centrifuged at 10,000 g followed by sequential washing with three wash buffers provided in the kit followed by centrifugation at 10,000g for 1 minute at each washing step. After complete drying of the spin column, the RNA was eluted out using a 50-100 μl elution buffer followed by centrifugation at 12,000 g for 1 minute.

### 2.4. Real Time (RT)-PCR analysis

Recently published Real Time-PCR assays were used for the detection of SARS-CoV-2 in wastewater samples (Ahmed et al., 2020; Corman et al., 2020). RT-PCR assays were performed using FDA approved Allplex™ 2019-nCoV Assay kit (cat# RP10244Y, RP10243X) or TaqPath™ COVID-19 Combo Kit (Cat#A47814) for the qualitative detection of SARS-CoV-2 genomic RNA in the sample on BioRAD CFX96 IVA Real-Time PCR and Applied Biosystems™ QuantStudio™ 5, respectively. The mastermix for Allplex™ 2019-nCoV Assay kit was prepared using the kit content which was composed of amplification and detection reagent, enzyme mix for one-time RT-PCR, buffer containing dNTPs, buffer for one-step PCR and RNase free water. Each PCR tube contained 8 μl RNA sample, 5μl 2019-nCoV MOM, 5 μl Real-time One-step buffer and 2 μl Real-time One-step enzyme and the final volume of the mixture was adjusted to 25μl using RNase free water. A list of different genes and fluorophores used for detection is given in Table 2. Thermal cycling reactions were performed at 50°C for 20 minutes, 95 °C for 15 minutes, 44 cycles at 94 °C for 15 seconds, and 45 cycles at 58 °C for 30 seconds, in a thermal cycler. For each run, a set of positive and negative controls were included. For the mastermix for TaqPath™ COVID-19 Combo Kit, each PCR tube contained 5μl RNA sample, 1.25 μl COVID-19 Real-Time PCR Assay Multiplex and 6.25μl TaqPath™ 1-Step Multiplex Master Mix (No ROX™) (4X) and the final volume of the mixture was adjusted to 25μl using RNase free water. Thermal cycling reactions began with UNG incubation at 25°C for 2 minutes, followed by reverse transcription at 53°C for 10 minutes, activation at 95°C for 2 minutes and 40 cycles of denaturation and annealing/ extension at 95°C for 3 seconds and 60°C for 30 seconds, respectively.

**Table 2:**
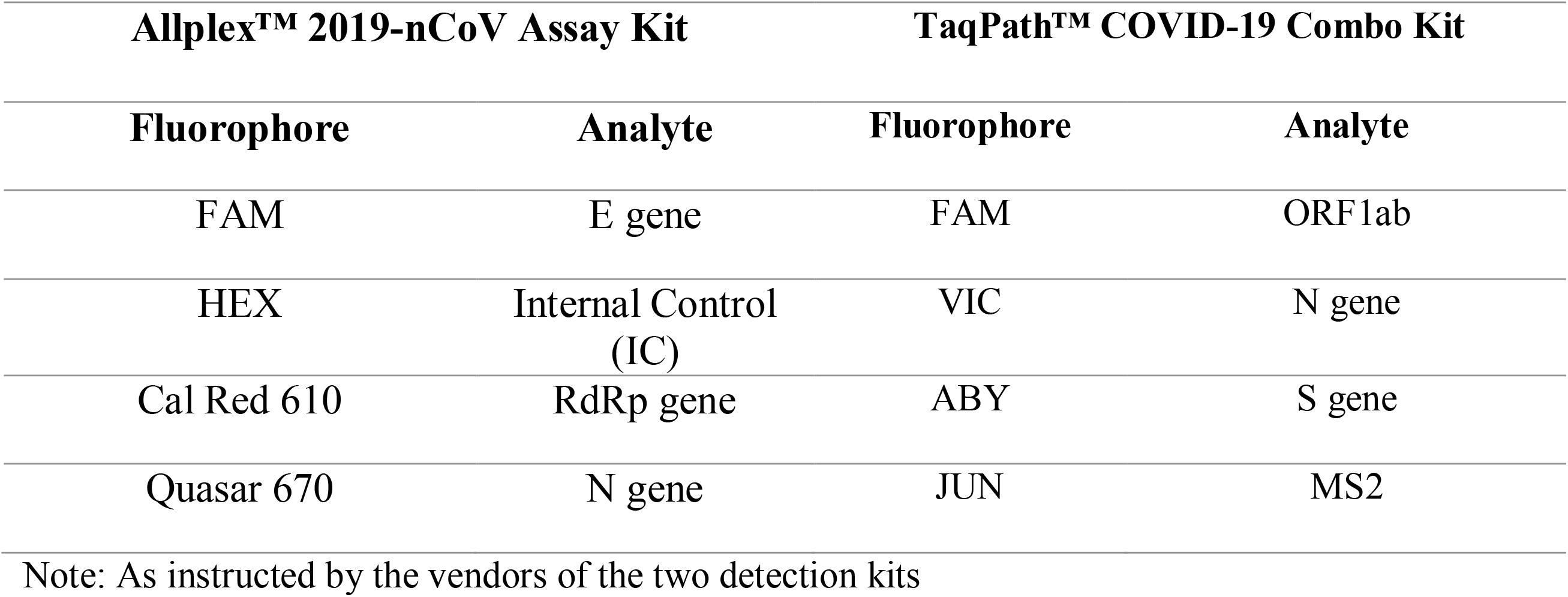
List of Fluorophores used for analyte detection in RT-PCR

## 3. Results

### 3.1. Detection of SARS-CoV-2 genome in wastewater samples

Six municipal WWTPs were selected from the Jaipur city, as shown in Table 1. The RT-PCR based detection was done in combinations of the following: one non-structural gene ORF1ab, any two of the structural genes like S (spike) or E (envelope) along with a third structural N (nucleocapsid) gene. COVID-19 RdRp (RNA dependent Replicase) was also used as a target to detect the presence of the COVID-19 genome with these structural genes. All the reactions were done with proper internal and positive controls followed by data analysis and result interpretation which is shown in Figure 2. A Ct value of <40 with Allplex kit and Ct values of <35 and <37 (for N gene and ORF1ab, respectively) with the TaqPath kit was considered to be valid and overall, if a sample showed two out of three genes with valid Ct values were considered to be overall positive for the presence of intact viral genome. The results of the study showed the presence of at least two target genes in untreated wastewater samples from site 5 and site 6 (as seen in Figure 3e & 3f), thus confirming the presence of SARS-CoV-2 genome in these samples. However, the other four wastewater samples tested negative for the detectable presence of the viral genome (Figure 3 a-d). The study tried to establish the correlations between the positively tested wastewater samples to the officially published public health data, of positive patients’ cases near the WWTPs during the study period. It was found that areas served by the WWTPs that showed positive results, reported a continuous increase in confirmed positive patients, soon after the first sampling. These observations were in accordance with the previous studies (Medema et al., 2020; Ahmed et al., 2020). The Ramniwas garden WWTP (Site 5) is currently serving the walled city area of Jaipur that includes the major hotspot of the city, the Ramganj area, having maximum reported cases.

**Figure 2:**
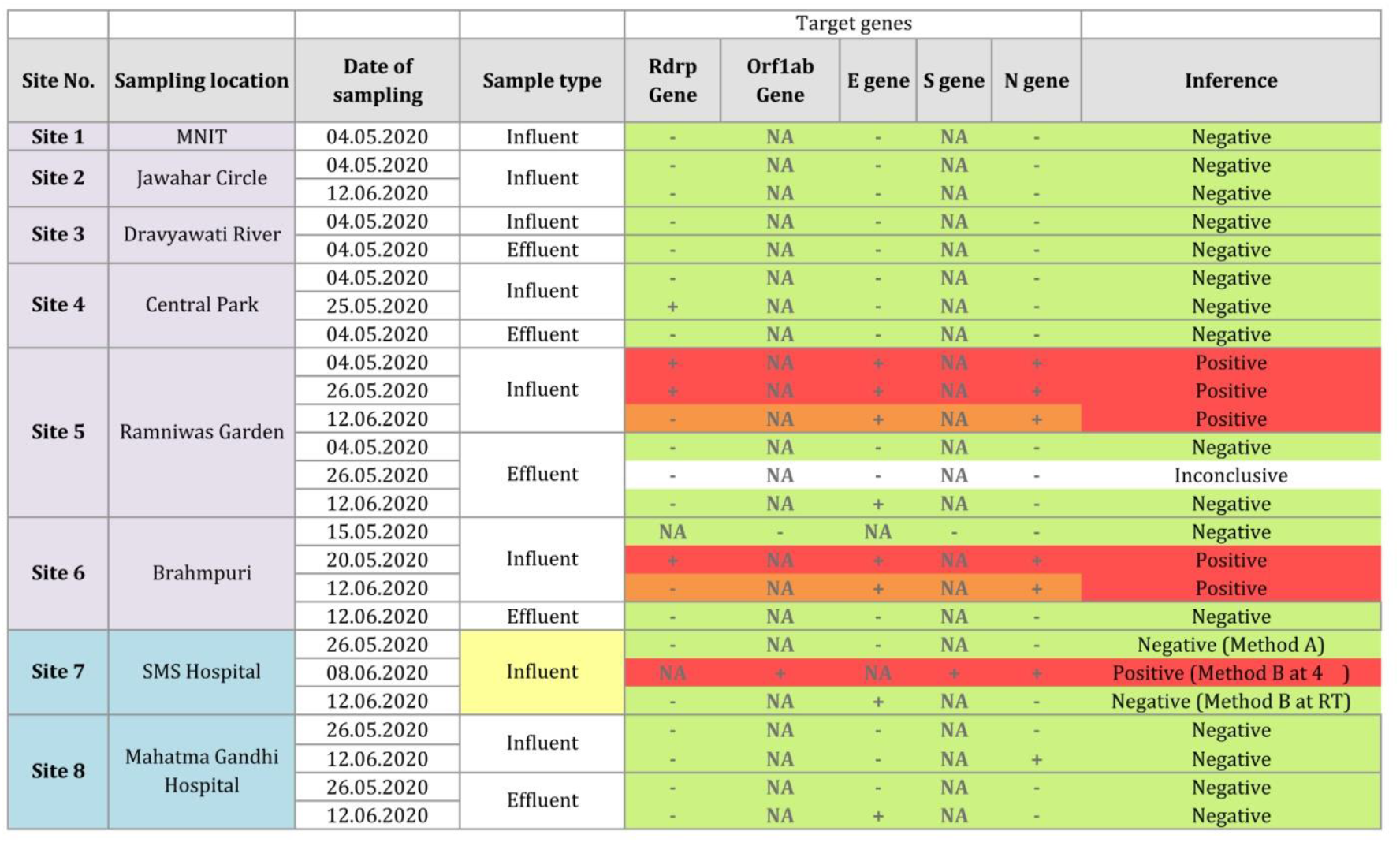
Comprehensive data analysis of sampling sites and result interpretation: The samples inferred positive are coded red while those inferred as negative sample for SARS-CoV-2 are coded in green. Samples which showed valid Ct for at least two of the three target genes are shown here in orange (as positive) while the samples with all the three genes having valid Ct values (as positive) are shown here in red. Site 7 (Influent) is color coded yellow, is discussed in section 3.5. Site 7, effluent sample, Inconclusive (marked white) shows the reaction for which internal control failed in RT-PCR detection.

**Figure 3:**
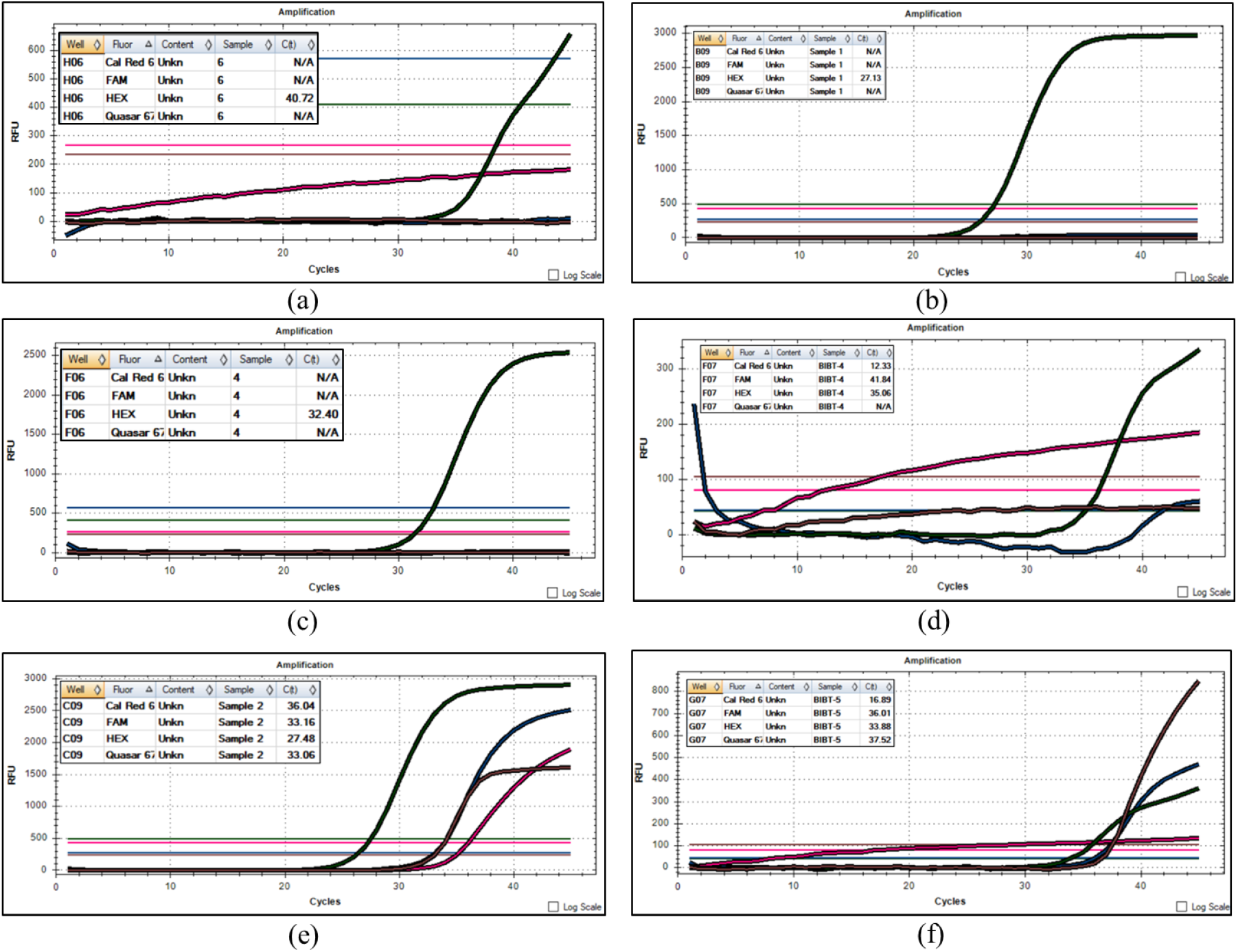
Results analysis of untreated wastewater samples collected from municipal WWTPs: The Graphs of Ct values were analyzed to determine positive and negative samples based on presence of the genome. The C_t_ values of <40 were considered to be valid for genes RdRp, E and N tested by Allplex kit. Those samples which had two out of three genes with valid Ct values were considered to be overall positive for the presence of intact viral genome. (a) Site 1, untreated wastewater sample showing negative results. (b) Site 2, untreated wastewater sample showing negative results (c) Site 3, untreated wastewater samples showing negative results (d) Site 4, untreated wastewater sample, showing negative results. (e) Site 5,untreated wastewater sample, showing all three genes tested (RdRp, E and N gene) had valid Ct values. (e) Site 6, untreated wastewater sample, showing all three genes tested (RdRp, E and N gene) had valid Ct values.

Our study reveals that a significant increase in the numbers of positive tested cases was reported within 6-14 days of our first date of sampling (May 4^th^, 2020). This increase was even more significant owing to a particular hotspot area of Jaipur city (Central jail), which showed a surge in the number of positive cases within six days of testing of the sample from site 5 (WWTP) that confirmed the presence of the virus.

Additionally, the untreated wastewater sample from Brahmpuri WWTP (site 6), the first sample (collected on 15^th^ May 2020) was interpreted negatively as it did not show the presence of virus, which correlated with the corresponding low number of positive cases around the area. However during sampling in the subsequent week, the Brahmpuri WWTP showed positive detection of the viral genome, and this was well correlated by a sudden surge in COVID-19 cases up to fivefold within the next six days of the detection. Previous reports have mentioned that the presence of the viral genome can be as early as 14 days of actual positive testing (Bernard Stoecklin et al., 2020). This difference in the gap of detection can be due to the following reasons: Firstly, in contrast to the previous studies, our window of testing started much later, when a complete lockdown was underway and the corona spread had already reached the third phase. Secondly, we also noticed that after 15^th^ May 2020, the numbers of COVID-19 positive cases increased and the gap between our detection and surge of positive tested cases of an area decreased from 14 to even 6 days. We acknowledge that in each of these areas where samples were tested positive, the number of officially reported positive cases by the city was only in double digits. This low number of cases reported by the city can be attributed to a very low number of COVID-19 tests done per area per day by the city. This data has reinforced our confidence in proposing that WWTPs can indeed serve as good checkpoints in predicting the surge in COVID-19 cases well ahead of the individual testing. This is critical as even a delay of three days in COVID-19 detection can lead to a potential spread to thousands of people (Mallapaty, 2020).

WBE serves as an important tool to trace the circulation of viruses in a community, providing opportunities to estimate their prevalence, and geographic distribution (Sinclair et al., 2008). Wastewater systems offer a practical approach to detect viruses excreted in the feces of an entire region. Using this approach it becomes possible to monitor the epidemiology of virus infections even if they are not evident by clinical surveillance, especially because traditional epidemiological approaches may be limited by the asymptomatic nature of viral infections and under diagnosis of clinical cases. SARS-CoV-2 is known to cause symptomatic or pauci-symptomatic infections (Nicastri et al., 2020), making it difficult to determine the actual degree of viral circulation in a community and in making comparisons among different countries that have different clinical diagnostic testing capabilities with even different methodology or assays. Meanwhile, wastewater surveillance could provide an unbiased method of evaluating the spread of infections in different areas, even where resources for clinical diagnosis are limited and when reporting systems are unavailable or not feasible, such as in many developing countries (Daughton, 2020). This approach allows comparisons between different geographic regions and assessments of the infected patients in an area. Our study also highlighted the importance of the WBE tool by monitoring viral RNA in wastewater to assess disease prevalence and spread in defined populations, which may prove beneficial for predicting COVID-19 related public health policy.

### 3.2. Validation of SARS-CoV-2 genome in wastewater samples by two different protocols

There has been growing evidence of the presence of the SARS-CoV-2 genome in sewage samples, but the major challenge in the detection is the lack of an optimized and standardized protocol (Bivins et al., 2020). As there are several reports for viral RNA isolation to check for the genomic presence of the COVID-19 causing virus in human fecal samples and sewage samples (Chen et al., 2020; Hindson, 2020; Wang et al., 2020a), the present study tried to establish a protocol, which would be robust under local conditions. As mentioned in section 2.2, both methods (A and B) were standardized to achieve the detection of viral RNA in the sample. In the first method (Method A), attempts were made to standardize a protocol for the samples with low expected viral load. In this protocol, as mentioned in Section 2.2, the samples were subjected to heat inactivation of virus particles. After the inactivation, the COVID-19 viral particles present in the sample were concentrated by filtration and adsorption onto PEG into 1 ml of final volume. This ensured an increased concentration of viral particles present per ml. Secondly, in method B, the study was able to establish a direct isolation method from the wastewater samples. This method was standardized keeping in mind the untreated wastewater samples and fecal samples where the probability of striking a heavy load of viral particles existed. In the direct method, the samples were subjected to a centrifugation step to remove larger solid particles and debris and directly processed for the RNA extraction and RT-PCR detection assay. The results of the study showed that the samples could be similarly tested for the detection of viral genomic RNA regardless of the pre-processing and concentration differences due to the two protocols. Thus, the samples from a particular site (processed by method A or B) tested either positive or negative regardless of the methods used and that the testing results were linked to the area, rather than the sample processing method.

These observations consolidated the results from previous studies that a regular testing of samples from WWTPs can potentially indicate the presence of coronavirus patients in the community. Additionally, the present study would try to establish a standardized protocol & methodology in the current scenario. The study is in accordance with the recent finding that wastewater-based epidemiology (WBE) can be used to detect and manage infectious disease transmission in communities (Nghiem, L. D., 2020). As many research groups across the globe are mobilizing to monitor wastewater for SARS-CoV-2 RNA for this purpose, it is necessary to validate methodologies, and data sharing to maximize the yield of WBE. Efficient harmonization of sampling, quality control, analysis methods and publications will help to ensure a high-quality evaluation of WBE (Bivins et al., 2020).

### 3.3. Detection of SARS-CoV-2 genome at high ambient temperatures during summers

As mentioned previously, wastewater treatment plants cater to tens of thousands of people in a given community. It has been previously reported that the virus can stay for very long periods in water and wastewater but at the same time, there are other studies that have reported that high temperatures can negatively affect the viral persistence outside the human body (La Rosa et al., 2020). In India especially in Rajasthan, however, where the temperatures can go as high as 45-50 °C in May-June, the study aimed to check if the WWTP samples could still have the presence of SARS-CoV-2 genome at higher temperature. The idea was to check if WWTPs in India could act as a checkpoint and alert against the COVID-19 outbreaks. Therefore, the samples were collected in the months of May and June 2020, where the ambient temperatures in the city were high up to 45°C. The presence of the viral genome was detected at a high ambient temperature of 45°C, which showed that WWTP samples can serve as a checkpoint where the presence of the COVID-19 genome can be checked. These results correlated with a recent study that predicted the linear upward (increasing) trend of positive patients been found in most states of India with increasing temperature and humidity (Goswami et al., 2020, Méndez-Arriaga, F. (2020).

### 3.4. Efficacy of Current Treatment systems at WWTPs against SARS-CoV-2

In India and other developing countries, the effluents from wastewater treatment plants find their way to the nearby gardens and agricultural areas for irrigation reuse. In this context, it becomes necessary to validate the presence of the viral genome in the treated effluent samples from WWTPs. It is highly likely that this wastewater may be originating from areas under coronavirus spread. It has been reported previously that the virus can persist with its active state (La Rosa et al., 2020) and this could pose a risk to the safety of people involved in the irrigation and public health in general. The contamination due to this wastewater may also lead to a new chain of viral spread, especially across the rural areas of India where medical facilities are limited. So, the present study also tried to investigate the efficacy of the current wastewater treatment technologies against COVID-19. Interestingly, the results of the study (as shown in Figure 4) revealed that all the treated wastewater samples showed negative results for the presence of viral RNA from all the WWTPs. From the present study, it was inferred that even in those WWTPs which tested positive for untreated wastewater; the treated wastewater had no detectable viral genome. This is important to mention that the positive site 5 (Ramniwas garden) currently uses Moving bed biofilm reactor (MBBR) technology, and site 6 (Brahmpuri) used Sequencing batch Reactor (SBR) technology, and the results conclude that the different wastewater treatments technologies being used are currently effective and able to decrease the viral particles below the detection limit, thus confirming the efficacy of treatment technology in attenuation of the virus, concluding that there is little risk to public health with the reuse of treated water for irrigation.

**Figure 4:**
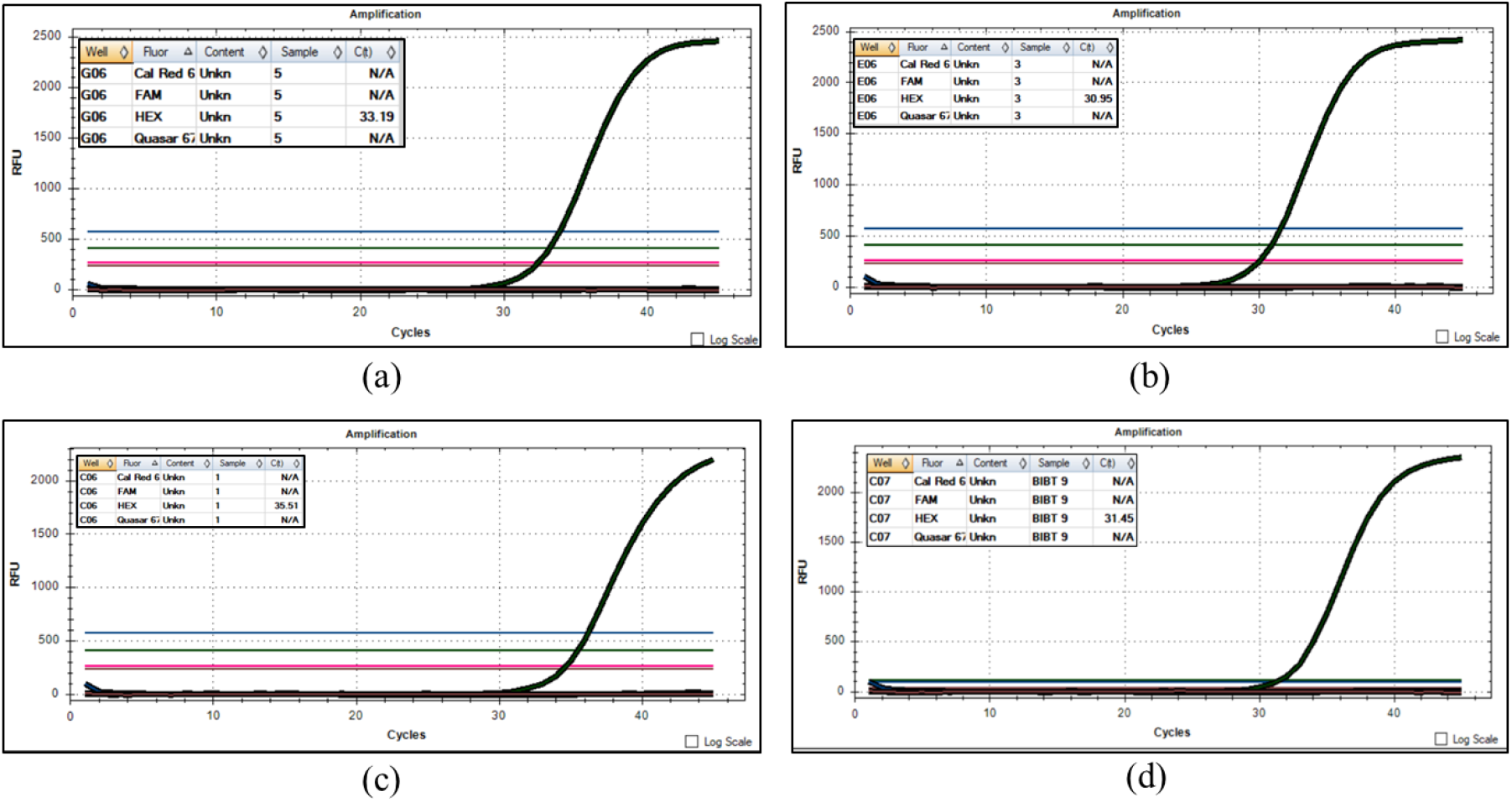
Results analysis of treated wastewater samples collected from the municipal WWTPs. *(a) Site 3 (b) Site 4 (c) Site 5 (d) Site 6*, shown here are graphs for Ct values for the three target genes tested. The graphs shown here have valid Ct value for the internal control but not for the target genes; hence all the samples are negative.

### 3.5. Efficacy of current sanitization practices (hypochlorite and other chlorine agents as disinfecting agent) in Hospitals

The present study also investigated decontamination and sanitization practices used in the hospitals. This is a critical aspect as although the suspected patients are quarantined from the general public, the waste they generate can potentially cause contamination to the external water resources after being discharged outside. The present study tried to investigate if there is any presence of the viral genomic RNA in the wastewater samples of two such local hospitals where patients with positive COVID-19 were being treated on campus. The results showed a discrepancy between the samples processed under the two different conditions, as highlighted in Figure 2 (coded yellow). The untreated wastewater samples of site 7, when processed by method A (using heat-inactivation and concentration methods) tested negative for the presence of viral genome. These results suggested that the heavy use of hypochlorite and detergent solutions used in current treatment systems in hospitals for sanitizing are effective in inactivating the viral genome and hence showed negative results (Wang et al., 2020b). The sanitization treatment seems to be successful in the de-stabilization of the viral coat in the case of enveloped viruses like COVID-19 that leads to faster degradation of the viral genomic RNA to below detection limits. Additionally, to further confirm our hypothesis, samples from one of the hospitals (site 7) was collected on 8^th^ June at 4°C and processed by direct protocol (Method B) and showed positive results (Figure 5) for the viral RNA. This confirms that this degradation was stalled by using lower temperatures for stabilizing coronavirus, as reported by La Rosa et al., 2020. The sample from site 7 was again collected on 12^th^ June, at ambient temperature and processed by Method B, interestingly gave negative results. As we know, the viral genome is possibly detected by the RT-PCR based tests, being sensitive enough to pick up even a single copy of the genome, this further confirmed our hypothesis. Thus, it highly likely to report that under the normal physiological conditions, the sanitization practices being followed by hospitals are able to removing the viral particles of COVID-19, and are safe to be reused or discharged in the public domain. The wastewater samples from the second hospital which we tested (Site 8), showed consistently negative results.

**Figure 5:**
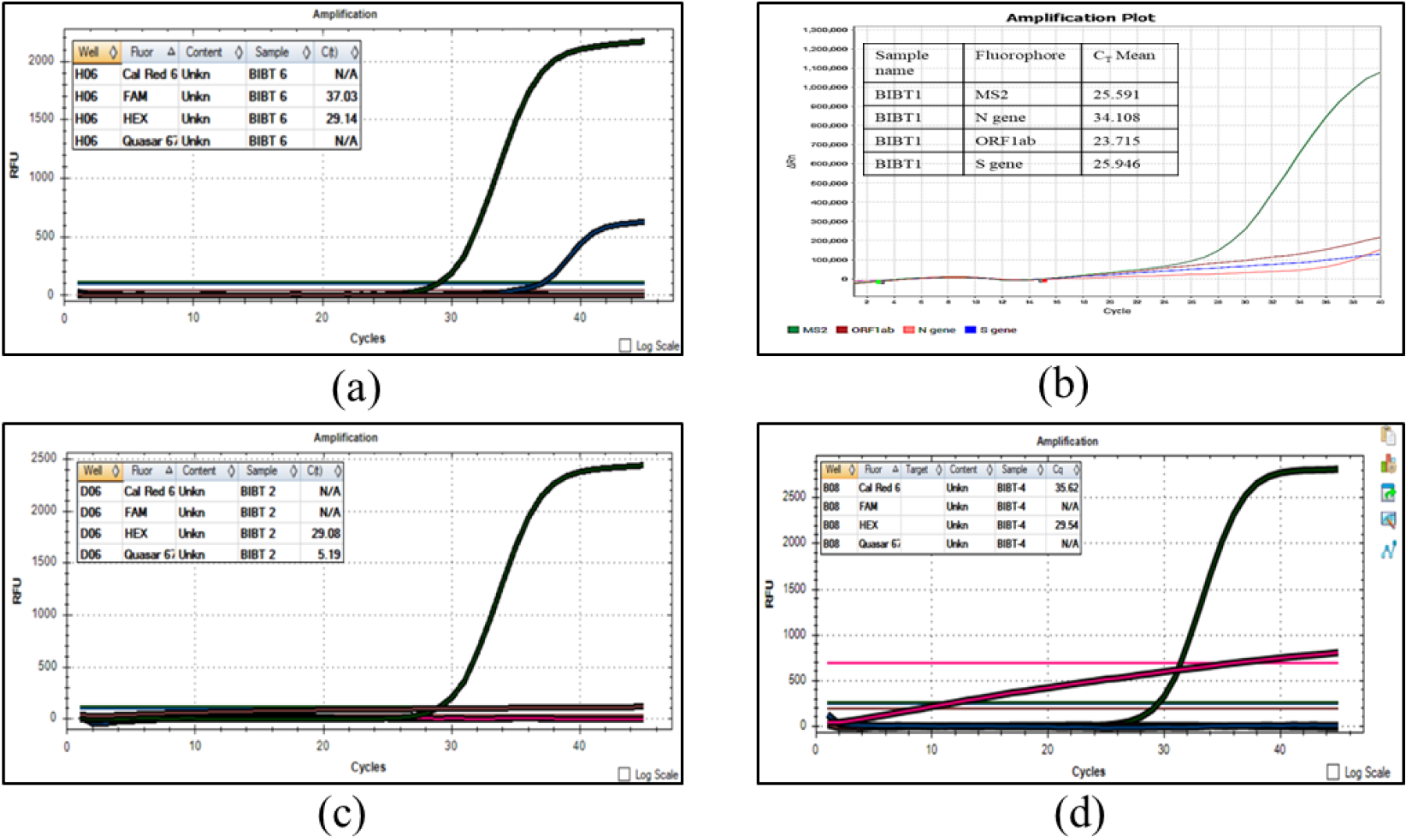
Results analysis of wastewater samples collected from the hospitals. (a) Site 7, untreated wastewater sample showing valid Ct for one gene (E gene) (processed at room temperature) (b) Site 7, untreated wastewater sample, showing all three genes tested (ORF1ab, S and N) had valid Ct values at 4°C. (c) Site 8, untreated wastewater samples showing negative results (d) Site 8, treated wastewater sample, showing negative results.

## 4. Discussions

Wastewater epidemiology is a useful study tool in monitoring any infectious disease spread and its community level dynamics in principle (Ahmed et al., 2020; Sims & Kasprzyk-Hordern, 2020). A previous study conducted in Paris demonstrated the detection of the viral genome before the exponential phase of the epidemic (Wurtzer et al., 2020), and another study by Randazzo et al., 2020 indicated that the SARS-CoV-2 can be detected weeks before the first confirmed case. The present study reports the first evidence of SARS-CoV-2 genome in wastewater samples from India, under higher ambient temperature conditions. Out of the municipal WWTPs that were selected for the current study, wastewater samples from two WWTPs (site 5 and 6) showed positive viral detection. We wondered if this pattern held any significance and if it could be related to the status of the positive tested cases in the respective area. We thus surveyed the officially published daily public health data of the total and new COVID-19 positive cases from the areas linked with our test WWTPs. The results of our study predicted that samples collected early from 4^th^ May to 15^th^ May, 2020, were tested positive, 10-14 days preceding a large jump in the number of reported corona positive cases in the respective area. However, during the late sampling (14th May to 12^th^ June, 2020), this gap decreased from 14 days to 6 days, between our detection and the surge of numbers in positive cases. One of the reasons for this contrasting trend might be the window of our observation. While the previous studies were done during the initial phases of disease spread along with the increasing imposition of restricted interactions in the respective countries, we have investigated the samples along with the progressive relaxation in the lockdown of the city. This is an interesting observation, and is reported for the first time. We thus hypothesize that this reduction in the gap could be because of a rapid spread of disease along with the accompanying relaxations in the lockdown in the city by the local government. Since, during a rapidly spreading pandemic, every day can potentially account for spread to tens of thousands of more people, we infer that this study highlights that a warning obtained even six days ahead, could be crucial in taking relevant steps against the outbreak.

The early detection of SARS-CoV-2 RNA in wastewater could have alerted about the imminent danger, giving a valuable time to the managers to coordinate and implement actions to curb the spread of the disease. Therefore, our outcomes support the proposition that WBE could be used as an early warning tool to monitor the status of COVID-19 infection within a community. Additionally, we believe that this environmental surveillance could be used as an instrument to drive the right decisions to reduce the risk of lifting restrictions too early. For instance, a key question is how to reduce the risk of a “second wave” and/or recurring local outbreaks. Massive population tests are the first choice, but in their absence, wastewater monitoring of SARS-CoV-2 RNA can give a reliable picture of the current situation.

The present study reports the evident presence of viral genome in untreated wastewater even during high ambient temperatures of 40-45°C. There have been epidemiology based studies outside India on COVID-19 virus which were done at colder seasons and quite low ambient temperatures (Bernard Stoecklin et al., 2020; Medema et al., 2020; Sims & Kasprzyk-Hordern, 2020). The detection of the COVID-19 genome in wastewater by these studies can be easily explained as the low range of temperature is reported to increase the duration of viral persistence (La Rosa et al., 2020). Our study also intended to investigate the feasibility of using the WBE approach yearlong for a country like India where temperatures can go as high as 50 °C. It is worth noting that, there are still no studies that report the viral behavior during high temperatures. This is the first study to check untreated and treated wastewater samples during the months of May-June, 2020 which are the hottest months in the summer especially in Jaipur (India). Thus, it is proposed that the testing of wastewater seems to be a useful approach in early detection and studying infection spread dynamics of the community population throughout the year.

In order to detect the presence of the viral genome, we have standardized two methods of sample processing. Firstly, Method A uses filtration and PEG adsorption method to increase the concentration of the SARS-CoV-2 virus before RNA extraction. This process was developed with the purpose of investigating probable samples with diluted loads. Secondly, Method B directly processes the sample for RNA extraction after removal of large suspended solids by a simple centrifugation. The current study tried to standardize Method B for the samples where we might get a relatively higher load of viral genome. However, we observed that both methods were effective uniformly in the case of municipal wastewater samples (sites 1 to 6), and we could detect the presence of SARS-CoV-2 in a sample regardless of the method employed. Thus each method acted as an internal control of another for each sample tested by them.

The most common use of the treated effluents from the WWTPs, is irrigation and it is imperative to check if the treated wastewater from the WWTPs, especially from the ones serving local hospitals, is safe to be used by the public in general. We, therefore, tested for the presence of COVID-19 genome in the treated wastewater samples of domestic as well as local public and private hospitals. Our results showed that the probability of the presence of any detectable viral genome load was very less and that the water could be considered safe for public consumption by the current standards. Since the hospitals we used for sample collections were following the sanitization protocol guidelines given by the ministry of health, we could conclude that the sanitization guidelines are sufficient to curb the viral presence and prevent the spread of this virus to a great extent in the present scenario. These kinds of studies are equally important to keep in check the sanitization practices being followed by Hospitals and in generating the new standard and guidelines, if required. The future implications of the work may include understanding the WBE tool as an alert system for preventing outbreaks.

## 5. Conclusions

- This is the first study that reports the detection of SARS-CoV-2 in wastewater in India using RT-PCR assay for the detection of viral genome in wastewater.
- The study also highlights the need for WBE tools and surveillance as an alert system that provides population-level estimates of the burden of SARS-CoV-2 against future outbreaks.
- This approach can become the basis for further developing a useful warning system for the cities of India, where in-person testing may not be available.
- The findings of the present study confirmed the detection of COVID-19 genome at ambient temperature (above 40°C) and can prove to be a useful tracking tool in understanding the dynamic behavior of this pandemic’s spread.
- The study highlights the efficacy of sodium hypochlorite or other chlorine compounds being used by hospital authorities as an effective disinfecting agent, to inactivate or attenuate viruses. However future studies would be required to further validate the virulence or infectivity of these viruses in wastewater samples.

## Data Availability

Authors will submit the data, as & when required.

## Credit authorship contribution statement

**Sudipti Arora**: Conceptualization, Investigation, Resources, Sample collection, Manuscript Writing and editing, corresponding author

**Aditi Nag**: Experimental design, protocol standardization, Experimental conduction, Data analysis, Writing -original draft of manuscript, Investigation

**Jasmine Sethi:** Experimentation, Pre processing, Manuscript preparation

**Jayana Rajvanshi**- Sampling, Experimentation, RT-PCR testing

**Sonika Saxena:** Resources providing, Sample collection protocol standardization and supervision

**Sandeep K. Shrivastava**- Experimental design in COVID-19 detection

**A. B. Gupta**- Conceptualization, Motivation, Supervision, Data analysis, Resource providing and manuscript review

## Declaration of competing interest

The authors declare that they have no known competing financial interests or personal relationship that could have appeared to influence the work reported in this paper.

## Funding

The present work was not funded by any external funding agency and all funds were incurred by the institute Dr. B. Lal Institute of Biotechnology, Jaipur.

## Acknowledgements

The study group would like to acknowledge the constant support received from Dr. B. Lal Gupta (Director, Dr. B. Lal Institute of Biotechnology, Jaipur) and Dr. Aparna Datta for inspiring this research and providing daily motivations to work faster. The team further acknowledges the efforts made by Mr. Dinesh and Mr. Rajeev for assisting sample collection. We would like to mention the support received from Dr. Deepika Gupta in guiding us for RT-PCR testing, and acknowledge the support received from the officials and persons at WWTPs who assisted us in sample collection.

## Notes

### Competing Interest Statement

The authors have declared no competing interest.

### Author Declarations

The present research work has received ethical clearance from Institutional Ethics Committee (IEC). The work does not include clinical trials.

